# Clinical Suspicion of COVID-19 in Nursing Home residents: symptoms and mortality risk factors

**DOI:** 10.1101/2020.10.28.20221333

**Authors:** Jeanine J.S. Rutten, Anouk M. van Loon, Janine van Kooten, Laura W. van Buul, Karlijn J. Joling, Martin Smalbrugge, Cees M.P.M. Hertogh

## Abstract

**Objectives:** To describe symptomatology, mortality and risk factors for mortality in a large group of Dutch nursing home (NH) residents with clinically-suspected COVID-19 who were tested with a Reverse Transcription Polymerase Chain Reaction (RT-PCR) test.

**Design:** Prospective cohort study.

**Setting and participants:** Residents of Dutch NHs with clinically-suspected COVID-19 and who received RT-PCR test.

**Methods:** We collected data of NH residents with clinically-suspected COVID-19, via electronic health records between March 18^th^ and May 13^th^, 2020. Registration was performed on diagnostic status (confirmed (COVID-19+)/ruled out (COVID-19-)) and symptomatology (typical and atypical symptoms). Information on mortality and risk factors for mortality were extracted from usual care data.

**Results:** In our sample of residents with clinically-suspected COVID-19 (N=4007), COVID-19 was confirmed in 1538 residents (38%). Although, symptomatology overlapped between residents with COVID-19+ and COVID-19-, those with COVID-19+ were three times more likely to die within 30 days (hazard ratio (HR), 3·1; 95% CI, 2·7 to 3·6). Within this group, mortality was higher for men than for women (HR, 1·8; 95%, 1·5-2·2) and we observed a higher mortality for residents with dementia, reduced kidney function, and Parkinson’s Disease, even when corrected for age, gender, and comorbidities.

**Conclusions and implications:** About 40% of the residents with clinically-suspected COVID-19 actually had COVID-19, based on the RT-PCR test. Despite an overlap in symptomatology, mortality rate was three times higher for residents with COVID-19+. This emphasizes the importance of using low-threshold testing in NH residents which is an essential prerequisite to using limited personal protective equipment and isolation measures efficiently.

## Background

A few weeks after the first reported coronavirus case in the Netherlands on February 27^th^,^1^ reports emerged of COVID-19 cases in nursing homes (NH). Due to a lack of Reverse Transcription Polymerase Chain Reaction (RT-PCR) test facilities and the limited available personal protective equipment, isolation measures were used in NH residents with clinically-suspected but not necessarily confirmed COVID-19. By April, one-third of the Dutch NHs reported at least one resident with COVID-19,^2^ and the number of deaths in NHs doubled during the COVID-19 pandemic.^3^

It is evident that severe acute respiratory syndrome coronavirus 2 (SARS-CoV-2) acts differently in older adults compared with younger adults.^4^ First, older adults have been reported to develop signs and symptoms more often, after being in contact with SARS-CoV-2.^4, 5^ Next, there are also indications for atypical disease presentation in older adults with symptoms such as delirium and falls^6-8^, and even for asymptomatic presentation^9^. In addition, shortness of breath and a high respiratory rate are reported to be more common in older adults.^5^ Indisputably, older age is associated with a higher mortality.^4, 5, 10-12^ However, accurate knowledge is lacking about the spectrum of symptomatology of COVID-19, mortality and risk factors for mortality in NH residents. Furthermore, it is unknown to what extent COVID-19 can be distinguished based on symptomatology from other infections (such as influenza, other viruses and bacterial infections) in NH residents, and whether a RT-PCR test is always necessary to make the distinction.

From March 18^th^ onwards, we set-up a COVID-19 registration, via an electronic health record (EHR) used by more than half of the NHs in the Netherlands. Combining data from this registration with usual care data extracted from the same EHR, we conducted a cohort study to:

- describe symptomatology and analyze mortality in residents with a clinical suspicion of COVID-19 and compare within this group the residents with confirmed COVID-19 to those where COVID-19 was ruled out as assessed a RT-PCR test.
- analyze risk factors for mortality in residents with confirmed COVID-19.

## Methods

### tudy design and setting

We conducted a prospective cohort study in Dutch NHs using a COVID-19 registration form linked to the EHR Ysis. Ysis, managed by software developer GeriMedica, is the most widely used EHR in NHs in the Netherlands and provides pseudonymized data on over 61,000 out of a total of 115,000 residents.^13^

### Selection of study population

We included residents with a clinical suspicion of COVID-19, based on the physicians assessment and of whom we had the result of the RT-PCR test. We excluded residents if results of follow-up diagnostics were not (yet) available. The Dutch Association of Elderly Care Physicians (Verenso) created a guideline which describes when to consider COVID-19, and when to perform a RT-PCR test.^14^ Because of scarcity, until 26 March it was advised to use a PCR test only in NH residents with at least two of the following symptoms: fever or feverish feeling, coughing and shortness of breath. In addition, it was advised only to test at wards without proven cases of COVID-19. Other NH residents with clinically-suspected COVID-19 were treated as COVID-19 patients upfront. From 26 March onwards, it was advised to perform a RT-PCR test in NH residents with any possibly COVID-19 related symptoms. From the first week of April, it was advised to test all NH residents with clinically-suspected COVID-19, also at wards with proven cases of COVID-19.

### Data collection

The study started on March 18^th^, 2020 and is ongoing. For this article, we analyzed data up to May 13^th^. If “Covid” or “corona” were registered in the EHR Ysis, a standardized assessment form appeared automatically on the EHR for the physician to complete. This standardized assessment form contains questions about the presence of ‘typical symptoms’ (fever, coughing, and shortness of breath), ‘atypical symptoms’ (sore throat and delirium/confusion/drowsiness), other symptoms, oxygen saturation (normal or decreased) and body temperature. All the symptoms were determined and registered by the physician(s) caring for the resident.

For all subsequent entries in the EHR for participants included in our cohort, a follow-up form was presented. The follow-up form contained a categorical question on follow-up diagnostics (COVID-19 confirmed, COVID-19 ruled out, no test outcome yet available).

Age, gender, type of ward, mortality, date of death, and comorbidity were derived from usual care data in the EHR. Data on comorbidity was registered by the physician(s) in open text fields in the medical history. We selected comorbidities that were considered risk factors for severe illness or mortality with COVID-19 in the general population by the Dutch National Institute for Public Health and the Environment at that moment. These comorbidities: dementia (including other cognitive disorders), chronic respiratory diseases, chronic cardiovascular disease, cerebrovascular diseases, diabetes mellitus, reduced kidney function, and Parkinson’s disease were extracted from the usual care data by an extensive search using MATLAB (The Mathworks, Natick, MA, USA). Search entries for each comorbidity were formed by three medical doctors (MS, JvK and JR), taking into account multiple variations in the way of registration. For example the search entry for ‘reduced kidney function’ contained the following keywords: impaired renal/kidney function, renal/kidney insufficiency, renal/kidney failure, dialysis, nephropathy etc. The search entries for each comorbidity were programmed in MATLAB. MATLAB then automatically searched the open text fields in which the physicians registered the medical history. For the first 200 residents the automated search was compared to a manual coding, and the programmed coding scheme was adjusted accordingly. The usual care data, data from the standardized assessment form and follow-up form were linked and pseudonymized by GeriMedica (the software development company who developed and manages the EHR).

### Ethics

The Medical Ethics Committee of the Amsterdam Medical Center reviewed and approved the study protocol.

### Statistical analysis

Symptomatology of residents with a clinical suspicion of COVID-19 was analyzed descriptively and separate for residents with COVID-19 confirmed (COVID-19+) and residents where COVID-19 was ruled out (COVID-19-) as assessed with a RT-PCR test. We used t-tests for continuous variables and chi-square tests for categorical variables to compare demographic characteristics and symptomatology. Survival curves on 30 days-mortality were estimated based on the days between the date of clinically-suspected COVID-19 and the date of death using Kaplan Meier curves in both residents with COVID-19+ and COVID-19-. The mortality rate in residents with COVID-19+ and COVID-19-was compared using a Cox proportional hazard model. The association between demographic characteristics, clinical characteristics, and mortality rate was analyzed in residents with COVID-19+ using three models: model 1 unadjusted, model 2 included gender and age, model 3 included gender, age and comorbidities. All model covariates were selected *a priori* on the basis of clinical relevance or results of the unadjusted model 1. Results are presented with 95% confidence intervals and all reported P values are two-sided. To indicate the number of missing values, the N on which percentages were calculated, are reported. All analyses were performed with the use of the SPSS statistical package, version 20·0 (IBM, Illinois, US).

## Results

Between March 18^th^ and May 13^th^, 2020, a clinical suspicion of COVID-19 was registered for 5425 NH residents (see Figure 1). Due to the lack of (results of) follow-up diagnostics, 1418 residents were excluded. Follow-up diagnostics confirmed COVID-19 in 1538 of the 4007 residents (38%); COVID-19 was ruled out in 2469 (62%) residents.

**Figure 1.**
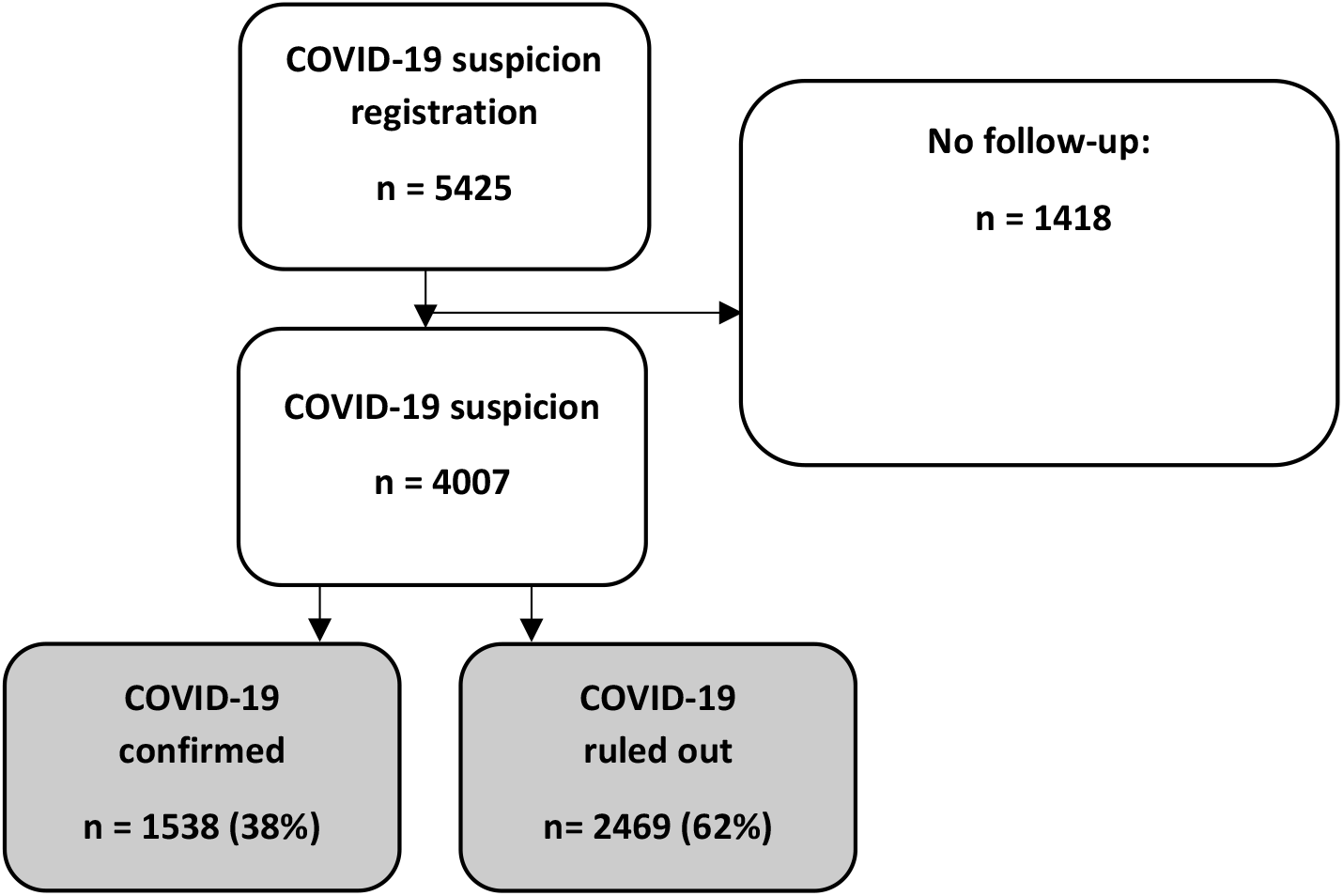
Flow-chart.

### Residents characteristics

Residents with a clinical suspicion of COVID-19 had a mean age of 84 years (SD: 9·8), were mostly women (62%) and resided mostly on psychogeriatric wards (42%) (Table 1). Residents with COVID-19+ were more likely to reside in a psychogeriatric ward (47% vs. 39%) and to have dementia (62% vs. 51%) and were less likely to have chronic respiratory disease (21% vs. 18%).

**Table 1.** Resident characteristics.

| **Table 1. Resident characteristics** |  |  |  |  |  |  |  |
| --- | --- | --- | --- | --- | --- | --- | --- |
|  | **COVID-19 confirmed**<br>**(N = 1538)** |  |  |  | **COVID-19**<br>**ruled out**<br>**(N = 2469)** | **Total study population**<br>**(N = 4007)** |  |
|  | **Men**<br>**(N =554)** | **Woman**<br>**(N = 984)** | **P-value**<br>**(men**<br>**versus**<br>**woman)** | **Total**<br>**(N =1538)** | **Total** | **P-value**<br>**(confirmed**<br>**versus**<br>**ruled out)** |  |
| Age<br>(mean, SD) | **82 (8.8)** | **85 (8.4)** | **<0.001** | **84 (8.7)** | **83 (10.4)** | **0.02** | **84 (9.8)** |
| Women | - | - | - | **64%** | **61%** | **0.07** | **62%** |
| **Ward type** |  |  |  |  |  |  |  |
| Psychogeriatric | **42%** | **50%** | **0.003** | **47%** | **39%** | **<0.001** | **42%** |
| Somatic | **18%** | **19%** | **0.81** | **19%** | **21%** | **0.12** | **20%** |
| Rehabilitation /<br>short term care | **18%** | **16%** | **0.35** | **16%** | **17%** | **0.48** | **17%** |
| Other | **1%** | **1%** | **0.46** | **1%** | **2%** | **0.25** | **2%** |
| Unknown | **21%** | **14%** | **0.001** | **17%** | **21%** | **0.001** | **19%** |
| **Comorbidity** |  |  |  |  |  |  |  |
|  | **N= 547\*** | **N=978\*** |  | **N=1525\*** | **N=2436\*** |  | **N = 3961\*** |
| Dementia | **58%** | **64%** | **0.02** | **62%** | **51%** | **<0.001** | **59%** |
| Cardiovascular<br>disease | **56%** | **47%** | **<0.001** | **50%** | **49%** | **0.28** | **53%** |
| Cerebrovascular<br>disease | **47%** | **38%** | **0.001** | **41%** | **41%** | **0.98** | **44%** |
| Diabetes<br>Mellitus | **25%** | **27%** | **0.53** | **26%** | **25%** | **0.28** | **27%** |
| Chronic<br>respiratory<br>disease | **20%** | **16%** | **0.09** | **18%** | **21%** | **0.02** | **21%** |
| Reduced kidney<br>function | **19%** | **18%** | **0.61** | **18%** | **19%** | **0.66** | **20%** |
| Parkinson<br>disease | **9%** | **5%** | **0.001** | **6%** | **7%** | **0.53** | **7%** |

\*Total number of residents with data on comorbidity.

Except for these differences, no important clinical differences were found between the characteristics of residents with COVID-19+ and COVID-19-. Of the 1538 residents with COVID-19+ 554 were men (36%). Compared with women, men were younger (82 vs. 85 years), and more often had cardiovascular disease (56% vs. 47%), cerebrovascular disease (47% vs. 38%), and Parkinson disease (9% vs. 5%). Women with COVID-19+ had dementia (64% vs. 58%) more often than men.

### Symptomatology

#### Typical symptoms

Most residents with COVID-19+ had one (42%) or two (35%) typical symptoms (i.e., cough, shortness of breath, fever); 9% did not have any typical symptoms.

Of the residents with COVID-19+, 63% presented with cough and 30% suffered from shortness of breath (Table 2). For residents where COVID-19 was ruled out this was 62% and 39%, respectively. Fever was more common in residents with COVID-19+ compared to residents where COVID-19 was ruled out (63% vs. 42%), whereas shortness of breath and sore throat were reported more frequently in residents with COVID-19- (39% vs. 30% and 13% vs. 10%).

**Table 2.** Symptomatology of NH residents with a clinical suspicion of COVID-19.

|  |  |  |  |  |  |  |
| --- | --- | --- | --- | --- | --- | --- |
|  | **COVID-19 confirmed** |  |  |  | **COVID-19 ruled out** |  |
|  | **Men**<br>**(N = 554)** | **Woman**<br>**(N = 984)** | **P-value**<br>**(men**<br>**versus**<br>**woman)** | **Total**<br>**(N = 1538)** | **P-value**<br>**(confirmed**<br>**versus**<br>**ruled out)** | **Total**<br>**(N = 2469)** |
| **Typical symptoms** |  |  |  |  |  |  |
| Cough | 62% (321/515) | 63% (579/916) | 0.74 | 63% (900/1431) | 0.75 | 62% (1440/2309) |
| Shortness of breath | 35% (174/498) | 28% (243/875) | 0.005 | 30% (417/1373) | <0.001 | 39% (882/2249) |
| Fever | 68% (361/533) | 60% (556/922) | 0.005 | 63% (917/1455) | <0.001 | 42% (969/2316) |
| **Atypical symptoms** |  |  |  |  |  |  |
| Sore throat | 8% (26/339) | 11%(68/637) | 0.13 | 10% (94/976) | 0.007 | 13% (233/1699) |
| Delirium/confusion<br>/drowsiness | 35%(170/482) | 25%(202/806) | <0.001 | 29% (372/1288) | 0.16 | 27% (570/2138) |
| **Other symptoms\*** |  |  |  |  |  |  |
| Fatigue | 19% (26/136) | 24% (67/281) | 0.28 | 22% (93/417) | <0.001 | 13% (112/869) |
| Diarrhea | 18%(24/136) | 18% (50/281) | 0.97 | 18% (74/417) | 0.43 | 16% (139/869) |
| Malaise | 17%(23/136) | 18%(50/281) | 0.82 | 18% (73/417) | 0.004 | 12% (101/869) |
| Rhinorrhea | 11%(15/136) | 13%(37/281) | 0.54 | 12% (52/417) | 0.23 | 15% (130/869) |
| Nausea/vomiting | 8%(11/136) | 13%(37/281) | 0.13 | 12% (48/417) | 0.001 | 19% (163/869) |
| Common cold | 14%(19/136) | 12%(33/281) | 0.52 | 12% (52/417) | 0.58 | 14% (118/869) |
| **Signs** |  |  |  |  |  |  |
| Decreased oxygen<br>saturation | 49%(187/383) | 42% (266/640) | 0.02 | 44% (453/1023) | 0.19 | 47% (820/1751) |
| Temperature |  |  |  |  |  |  |
| <36.5°C | 4% (17/450) | 5% (37/763) | 0.48 | 4% (54/1213) | <0.001 | 12% (220/1883) |
| 36.5 – 37.5 °C | 14% (64/450) | 18% (140/763) | 0.14 | 17% (204/1213) | <0.001 | 33% (613/1883) |
| 37.5 – 38.0 °C | 14% (64/450) | 18% (137/763) | 0.19 | 17% (201/1213) | 0.64 | 16% (310/1883) |
| >38.0 °C | 68% (305/450) | 59% (449/763) | <0.001 | 62% (754/1213) | <0.001 | 39% (740/1883) |

\*Not assessed via categorical questions, but reported in an open text field

#### Atypical and other symptoms

The risk of delirium/confusion/drowsiness and sore throat in residents with COVID-19+ was comparable to the incidence of these symptoms in residents with COVID-19- (29% and 10% vs 27% and 13%) (Table 2). Other symptoms, which could be outlined in an open text field, were reported for 417 (27%) residents with COVID-19+ and for 869 (35%) of residents with COVID-19-. Fatigue was most commonly reported in residents with COVID-19+ (22%), followed by diarrhea (18%), malaise (18%), rhinorrhea (12%), nausea/vomiting (12%), and common cold (12%) *(Table 2)*. Other symptoms such as headache and anosmia were reported in less than 10% of cases. Fatigue and malaise were reported more often in residents with COVID-19+ than in residents with COVID-19- (22% vs. 13% and 18% vs. 12%), whereas nausea/vomiting was reported less often in residents with COVID-19+ (12% vs. 19%).

#### Signs

Of the residents with COVID-19+, 44% had decreased oxygen saturation, vs. 47% of the residents with COVID-19-. Of the residents with COVID-19+, most had a temperature of >38 °C (62%) compared to the residents with COVID-19- (39%).

### Mortality in NH residents with a clinical suspicion of COVID-19

The median number of days on which NH residents with COVID-19 died was 22 days after the day of the COVID-19 suspicion (Interquartile range (IQR) = 21), vs. 28 (IQR = 15) for COVID-19. Of the residents with COVID-19+, 42% had died in 30-days (95% confidence interval [CI], 39 to 44%), vs. 15% of the residents with COVID-19- (95% CI, 14 to 17%) (see Figure 2A). Residents with COVID-19+ were three times more likely to die within 30 days than residents with COVID-19- (adjusted hazard ratio (HR), 3·14; 95% CI, 2·7 to 3·6; P<0·001).

**Figure 2.**
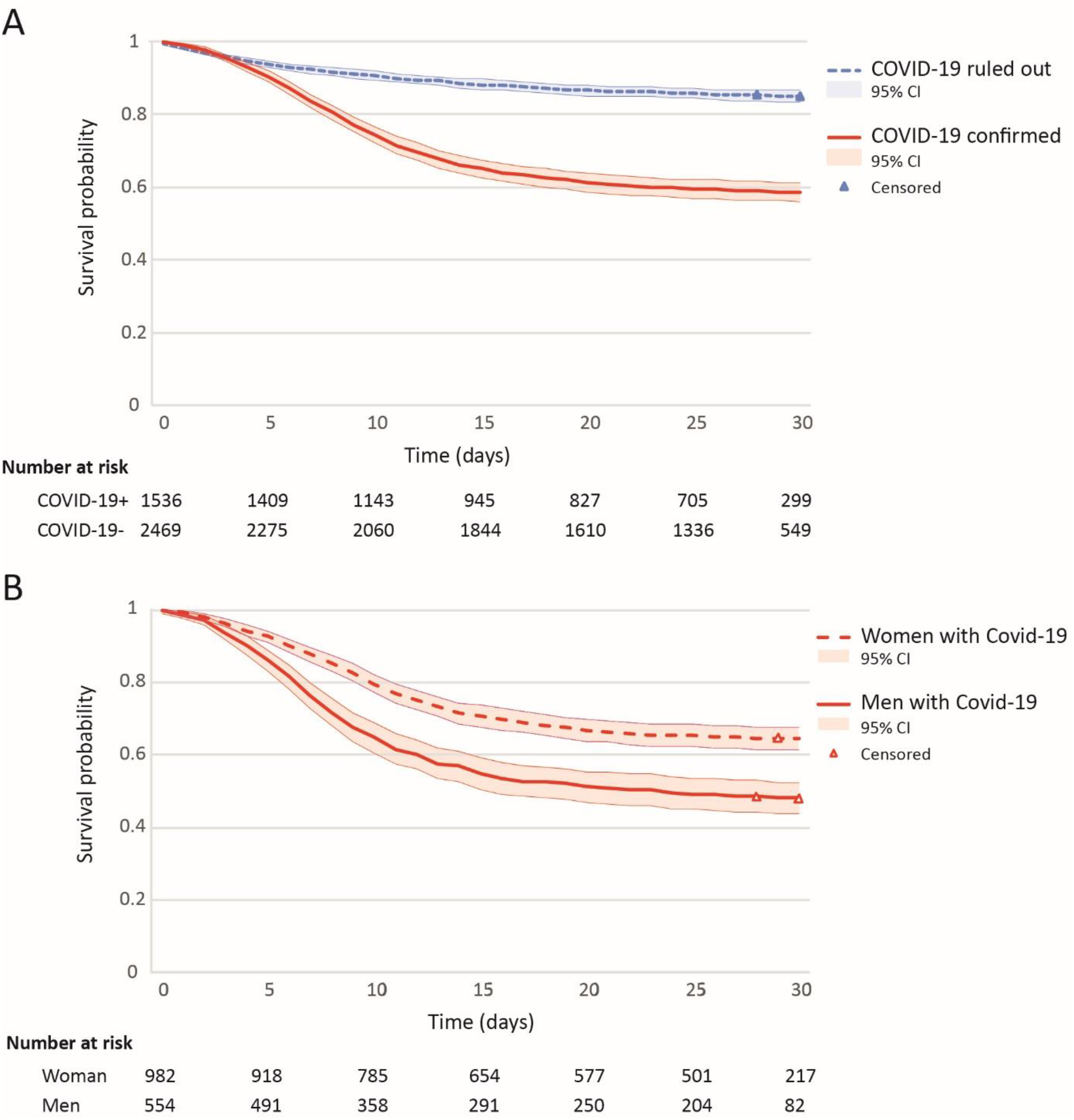
Kaplan-Meier estimates of survival for: **A: NH residents with confirmed and ruled out COVID-19**. The blue line indicates the 30-day survival probability of NH residents where COVID-19 was ruled out; the red line indicates the 30-day survival probability of NH residents with confirmed COVID-19. The triangles indicate censored data points. **B: men and women with confirmed COVID-19**. The dashed line indicates the 30-day survival probability of women with confirmed COVID-19; the solid line indicates the 30-day survival probability of men with confirmed COVID-19. The triangles indicate censored data points.

### Risk factors for mortality in residents with confirmed COVID-19

Gender, age, and comorbidity were associated with a higher mortality in residents with COVID-19+ (Table 3).

**Table 3.** Cox proportional Hazard model for risk factors for mortality in residents with confirmed COVID-19.

|  | **Model 1: unadjusted** |  | **Model 2: adjusted for gender and age** |  | **Model 3: adjusted for gender, age and relevant comorbidity†** |  |
| --- | --- | --- | --- | --- | --- | --- |
|  | HR (95% CI) | P-value | HR (95% CI) | P-value | HR (95% CI) | P-value |
| **Gender** |  |  |  |  |  |  |
| Women | 1.00 (ref) |  | 1.00 (ref) |  | 1.00 (ref) |  |
| Men | 1.70 (1.45-2.00)* | <0.001* | 1.84 (1.56-2.17)* | <0.001* | 1.82(1.54-2.15)* | <0.001* |
| **Age (years)** |  |  |  |  |  |  |
| = < 80 | 1.00 (ref) |  | 1.00 (ref) |  | 1.00 (ref) |  |
| 81-85 | 1.08 (0.85-1.37) | 0.55 | 1.12 (0.88-1.42) | 0.37 | 1.07 (0.84-1.37) | 0.57 |
| 86-90 | 1.37 (1.11-1.69)* | 0.004* | 1.56 (1.26-1.94)* | <0.001* | 1.49 (1.19-1.85)* | <0.001* |
| > 90 | 1.20 (0.96-1.51) | 0.11 | 1.41 (1.12-1.78)* | 0.004* | 1.38 (1.09-1.75)* | 0.008* |
| **Comorbidity** |  |  |  |  |  |  |
| Dementia | 1.23 (1.04-1.46)* | 0.02* | 1.25 (1.05-1.48)* | 0.01* | 1.26 (1.06-1.50)* | 0.008* |
| Chronic cardiovascular disease | 1.22 (1.04-1.43)* | 0.02* | 1.15 (0.98-1.36) | 0.08 | 1.15 (0.97-1.35) | 0.10 |
| Cerebrovascular disease | 1.10 (0.94-1.29) | 0.24 | 1.08 (0.92-1.27) | 0.33 | 1.06 (0.89-1.25) | 0.53 |
| Diabetes Mellitus | 0.99 (0.83-1.19) | 0.91 | 1.03 (0.86-1.23) | 0.79 | 0.99 (0.82-1.19) | 0.90 |
| Chronic respiratory disease | 0.97 (0.79-1.20) | 0.79 | 0.94 (0.76-1.16) | 0.55 | 0.92(0.75-1.14) | 0.46 |
| Reduced kidney function | 1.39 (1.15-1.69)* | 0.001* | 1.36 (1.13-1.65)* | 0.002* | 1.35 (1.11-1.64)* | 0.002* |
| Parkinson's disease | 1.44 (1.08-1.91)* | 0.01* | 1.40 (1.05-1.88)* | 0.02* | 1.49 (1.11-2.00)* | 0.007* |

† Relevant comorbidity includes dementia, chronic cardiovascular disease, reduced kidney function and Parkinson's disease

#### Gender-differences

In comparison with women, men were more likely to have experienced severe symptoms such as shortness of breath (35% vs. 28%), fever (68% vs. 60%), delirium/confusion/drowsiness (35% vs. 25%), and decreased oxygen saturation (49% vs. 42%) in the COVID-19+ group (Table 4).

Of men with COVID-19+ 52% had died within 30 days (95% CI, 48 to 56%), vs. 36% of women with COVID-19+ (95% CI, 33 to 39%). The median number of days, after the day of the COVID-19 suspicion, on which male NH residents with COVID-19 died was 16 days (IQR = 23), vs. 25 days (IQR = 9) for women with COVID-19. We found an HR of 1·8; 95% CI, 1·6 to 2·2; P<0·001 when adjusted for age and comorbidities (see Figure 2B).

#### Comorbidities

Dementia (HR 1·26; CI, 1·06-1·50; P= 0·008), reduced kidney function (HR 1·35; CI, 1·11-1·64; P=0·002), and Parkinson’s disease (HR 1·49; CI, 1·11-2·00; P=0·007) were associated with a higher mortality rate in residents with COVID-19+ when adjusted for age, gender, and comorbidities. For other comorbidities, no statistically significant associations were found.

## Discussion

COVID-19 was only confirmed in 1538 (38%) of the 4007 residents with a clinically-suspected COVID-19. Many NH residents with confirmed COVID-19 had ‘typical symptom(s)’ such as fever, cough, or shortness of breath. Atypical symptoms were also reported regularly (i.e. delirium/ confusion/drowsiness, fatigue, diarrhea, malaise, rhinorrhea, nausea/vomiting, and common cold). We observed an overlap in symptoms between residents with and without confirmed COVID-19 following clinically-suspected COVID-19. However, fever occurred more frequent in the COVID-19 confirmed group (63% versus 42%). Furthermore, the mortality rate in residents with confirmed COVID-19 was three times higher than in residents where COVID-19 was ruled out. For residents with confirmed COVID-19, the 30-day mortality in men was higher than in women. In addition, dementia, reduced kidney function, and Parkinson’s disease were all associated with a higher mortality rate in residents with confirmed COVID-19, even when corrected for age, gender, and comorbidities.

In our sample, 63% of the NH residents with confirmed COVID-19 presented with fever, 63% with cough and 30% with shortness of breath. This is in line with a previous study that showed that fever (75%) was most common in older adults, followed by cough (44%) and shortness of breath (25%).^11^ In contrast, another study in older people showed that cough (80%) was more common than fever (60%).^15^ However, the sample sizes of the age groups (> 80 years) was much smaller in both studies (N = 16 and N = 20). Our study showed that COVID-19 in NH residents presents itself with both typical and atypical symptoms.

In our study, almost half of the residents with confirmed COVID-19 (42%) died within 30 days. This percentage is higher than the 34% reported by McMichael et al.,^16^ and the 26% reported by Arons et al.^9^ A possible explanation could be that in our sample, in part because of the RT-PCR test shortage in the early weeks of patient enrollment, only residents with symptoms were tested, whereas in the abovementioned studies residents were also tested in the absence of symptoms. In comparison with asymptomatic presentation of COVID-19, symptomatic presentation will result in a higher mortality. Because of the high mortality in this vulnerable group, it is necessary to protect NH residents from COVID-19. Especially, now that the flu season has arrived and the virus may spread even faster.^17^

We observed that in residents with confirmed COVID-19, men were more likely to die than women. This difference in mortality has also been found in other studies on COVID-19,^10, 18^ and preliminary data indicate an association between comorbidities, such as cardiovascular disease, and severity of COVID-19.^5, 19^ However, we adjusted our survival analyses for comorbidities, yet the difference in mortality between men and women remained. It has been suggested that this gender difference might be related to gender differences in SARS-CoV-2 entry receptors.^20^ Nonetheless, it remains unclear whether increased mortality for men is specific to COVID-19. In general, men are sick more often and more severely sick due to gender differences in hormones, immune responses and immune aging, and gender-specific differences in lifestyle and health behavior.^21^ Therefore, the observed difference in COVID-19 mortality between men and woman may be explained by the men-women disability survival paradox.^22^ This is the phenomenon that higher life expectancy is accompanied by higher rates of disability in women compared with men, but disability increases risk of mortality more in men than women. Therefore, men might be less resilient to acute infectious diseases as COVID-19.

Furthermore, within our NH residents with confirmed COVID-19, we observed a higher mortality in those with dementia, reduced kidney function, and Parkinson’s Disease (PD) than those without these comorbidities, even when corrected for age, gender, and comorbidities. Similarly, Willamson et al. showed that dementia/stroke and kidney diseases are risk factors.^23^ In contrast to our study, they also found an association between diabetes mellitus, cardiovascular-, neurological-, and respiratory diseases and mortality. Prior to our study, PD had been suggested as a comorbid risk factor for COVID-19 mortality, but empirical data was lacking.^24^ We show, for the first time in a large study population, that PD increased mortality risk of COVID-19 in NH residents. One possibility for this is an increased chance of death arising from pneumonia in PD patients,^24, 25^ although further data is required to test this possibility.

This study had several limitations. Based on the Verenso guideline, we assumed that follow-up diagnostics consists of a RT-PCR test. A RT-PCR test has relatively low sensitivity (63-78%).^26^ Consequently it could be that the group of residents where COVID-19 was ruled-out partly consisted of residents who actually had COVID-19. Moreover, the group of residents where COVID-19 was ruled out did have symptoms, which also may have caused by other underlying infections such as influenza or other viruses. It would be useful to conduct an additional case-control study in which a cohort of residents with confirmed covid-19 would be compared with a pre-COVID-19 era cohort to compare the mortality outcomes.

Secondly, our study was set up in the very early stages of the COVID-19 outbreak in the Netherlands (18^th^ March, 2020) and our standardized assessment form was created with available knowledge at the time. As a result, not all relevant symptoms were listed in the standard form. However, we included a question about delirium/confusion/drowsiness in the standardized form, since we hypothesized that NH residents could have atypical presentations. Moreover, physicians had the opportunity to report other symptoms in an open text field, and did so for 32% of the participants in this study. Of note, because of the shortage of RT-PCR tests in the beginning of the epidemic in the Netherlands, initially only residents who met the criteria as described in the Verenso guidelines (i.e. residents with fever, cough, or shortness of breath) were tested. As a result, we were not able to describe the occurrence of an atypical or asymptomatic presentation of COVID-19 properly. A study in American NH residents found that almost half of residents with COVID-19 were asymptomatic at the time of testing, but developed symptoms later.^9^

## Conclusions and Implications

In a large cohort of clinically-suspected COVID-19 NH residents, 38% of the residents with clinically-suspected COVID-19 actually had a confirmed COVID-19. There was an overlap in symptoms between residents with confirmed COVID-19 and those for whom COVID-19 was ruled out, but we found that mortality rate in residents with confirmed COVID-19 was three times higher. This emphasizes the importance of using a low threshold to test NH residents for SARS-CoV-2. This is of vital importance since detecting positive COVID-19 diagnoses in NHs is extremely important, and an essential prerequisite to using limited personal protective equipment and quarantine and isolation measures efficiently. Furthermore, our data demonstrated that COVID-19 affects male residents disproportionally, mortality was almost twice as high for men than for women. It has been suggested that gender-differences in comorbidity could explain this effect. However, our data demonstrates that the increased mortality risk for men remained even when corrected for age and comorbidities.

## Data Availability

-

## Notes

### Competing Interest Statement

The authors have declared no competing interest.

### Funding Statement

Dutch Ministry of Health, Welfare and Sport, 329517

